# Estimating the early death toll of COVID-19 in the United States

**DOI:** 10.1101/2020.04.15.20066431

**Authors:** Daniel M. Weinberger, Ted Cohen, Forrest W. Crawford, Farzad Mostashari, Don Olson, Virginia E Pitzer, Nicholas G Reich, Marcus Russi, Lone Simonsen, Anne Watkins, Cecile Viboud

## Abstract

**Background:** Efforts to track the severity and public health impact of the novel coronavirus, COVID-19, in the US have been hampered by testing issues, reporting lags, and inconsistency between states.

Evaluating unexplained increases in deaths attributed to broad outcomes, such as pneumonia and influenza (P&I) or all causes, can provide a more complete and consistent picture of the burden caused by COVID-19.

**Methods:** We evaluated increases in the occurrence of deaths due to P&I above a seasonal baseline (adjusted for influenza activity) or due to any cause across the United States in February and March 2020. These estimates are compared with reported deaths due to COVID-19 and with testing data.

**Results:** There were notable increases in the rate of death due to P&I in February and March 2020. In a number of states, these deaths pre-dated increases in COVID-19 testing rates and were not counted in official records as related to COVID-19. There was substantial variability between states in the discrepancy between reported rates of death due to COVID-19 and the estimated burden of excess deaths due to P&I. The increase in all-cause deaths in New York and New Jersey is 1.5-3 times higher than the official tally of COVID-19 confirmed deaths or the estimated excess death due to P&I.

**Conclusions:** Excess P&I deaths provide a conservative estimate of COVID-19 burden and indicate that COVID-19-related deaths are missed in locations with inadequate testing or intense pandemic activity.

**RESEARCH IN CONTEXT:** *Evidence before this study:* Deaths due to the novel coronavirus, COVID-19, have been increasing sharply in the United States since mid-March. However, efforts to track the severity and public health impact of COIVD-19 in the US have been hampered by testing issues, reporting lags, and inconsistency between states. As a result, the reported number of deaths likely represents an underestimate of the true burden.

*Added Value of this study:* We evaluate increases in deaths due to pneumonia across the United States and relate these increases to the number of reported deaths due to COVID-19 in different states and evaluate the trajectories of these increases in relation to the volume of testing and to indicators of COVID-19 morbidity. This provides a more complete picture of mortality due to COVID-19 in the US and demonstrates how delays in testing led to many coronavirus deaths not being counted in certain states.

*Implications of all the available evidence:* The number of deaths reported to be due to COVID-19 represents just a fraction of the deaths linked to the pandemic. Monitoring trends in deaths due to pneumonia and all-causes provides a more complete picture of the tool of the disease.

## Introduction

The novel coronavirus SARS-CoV-2 first emerged in December 2019 in Wuhan, China and rapidly grew into a large-scale global pandemic.^1^ Tracking the severity and impact of COVID-19 is, at the time of writing, a critical need, hampered by testing issues and reporting lags for key epidemiological indicators.

Many countries, including the US, were caught off-guard by the speed with which COVID-19 spread from China. Without adequate capacity to test for the SARS-CoV-2 virus causing COVID-19 for much of February and March 2020, available laboratory-confirmed cases captured only an estimated 10-15% of all infections.^2^ Although most countries have adopted the strategy to preferentially test severe cases, estimating the number of severe infections and deaths caused by COVID-19 will be a challenge. Typically, a large proportion of deaths caused by infectious diseases are not attributed to a specific pathogen. With the limited availability of testing for the novel coronavirus and imperfect sensitivity of the tests,^3, 4^ there have undoubtedly been a number of deaths caused by the virus that are not counted in official tallies. Even in situations of ample testing, deaths from viral pathogens, including SARS-CoV-2, can occur indirectly via secondary bacterial infections or exacerbation of chronic conditions. Further, in the midst of a large outbreak, there is an unavoidable delay in compilation of death certificates and ascertainment of cause of deaths, which contributes to uncertainty about severity and burden.

Finally, in the US, there has been a high degree of variability in public health resources, laboratory testing, and recognition of the outbreak at the state level, which could lead to significant under-estimation of the true impact of the outbreak in certain geographies.

To estimate the burden of death due to novel respiratory pathogens, previous studies have compared the observed incidence of influenza-related deaths ascribed to pneumonia and influenza (“P&I”) with the baseline incidence of P&I that would be expected at that time of year.^5, 6^ These “excess deaths” provide an estimate of pathogen-specific burden. This approach was used in the early months of the 2009 influenza A/H1N1-pdm pandemic; it was estimated that just 1 in 7 pandemic-related deaths was captured by laboratory testing in 2009 in the US.^7, 8^

In this study, we estimated the increase in P&I deaths across the United States in each week in excess of a seasonal baseline to capture the direct and indirect mortality burden of COVID-19. We compare these estimates of excess deaths to the reported numbers of deaths due to COVID-19 in different states and evaluate the trajectories of these increases in relation to the volume of testing and to indicators of COVID-19 morbidity. These analyses provide insights into the burden of COVID-19 in the early months of the outbreak in the United States and serve as a surveillance platform that can be updated as new data accrue.

## METHODS

### Data

Data on deaths due to pneumonia and influenza (P&I, ICD-10 codes J09-J18) and all-causes by state and week were obtained from the National Center for Health Statistics’ (NCHS) mortality surveillance system.^9^ Here we analyze spikes in P&I mortality rather than straight pneumonia to be more comprehensive, as influenza-coded deaths do not necessarily require laboratory confirmation of influenza infection, there is overlap of symptoms between influenza and COVID-19, and P&I mortality has been used in US to monitor the severity of influenza and other respiratory pathogens since the 1918 pandemic. In addition, analysis of spikes in all-cause mortality provides a full picture of the direct and indirect burden of COVID-19.

The NCHS mortality data are available with a 2-week lag and are partially complete for the most recent weeks. The P&I mortality data provide the provisional number of deaths due to pneumonia or influenza (ie, deaths with a code of pneumonia or influenza anywhere in the death certificate) and should be adjusted by the total number of deaths reported in real time each week. Connecticut, North Carolina, and West Virginia were missing mortality data for recent months and were therefore excluded from the analyses.

The P&I grouping will include individuals who had a listed cause of death as COVID-19 (along with a P&I code) as well as people who did not have COVID-19 listed as a cause of death. It does not capture people who had COVID-19 listed as a cause of death but did not have P&I recorded among the causes.

We also compiled data on COVID-19-related morbidity to gauge the timing and intensity of the pandemic in different locations. Influenza-like illness (ILI) is a longstanding indicator of morbidity from acute respiratory pathogens, including SARS-CoV-2. Weekly state-level ILI data were obtained from the CDC’s ILINet system^10^, which aggregates data from a network of outpatient providers. To adjust for activity of non SARS-CoV-2 respiratory pathogens, we used state-level data on laboratory-confirmed influenza activity from the CDC’s National Respiratory and Enteric Virus Surveillance System (NREVSS)^11^. This dataset captures the number of tests performed for influenza and the number that were positive by week and state. The ILI data provide the percent of visits to participating outpatient providers that were for ILI. ILINet and NREVSS data are available with a 1-week lag.

The ILI, NREVSS, and mortality datasets were accessed through the CDC’s FluView portal using the cdcfluview package in R. Data from NCHS, ILINet and NREVSS were obtained for the weeks ending January 5, 2013 through March 28, 2020.

To compare our burden estimates with official COVID-19 tallies, we compiled weekly reports of laboratory-confirmed deaths due to COVID-19 in each state from several sources, including the Covid Tracking Project,^12^ and NCHS^13^. State-specific testing information was obtained was The Covid Tracking Project ^12^.

### Excess mortality and morbidity analysis

The 9 largest states by population, and Washington state, were analyzed individually. The remaining states for which we had data were grouped into the corresponding health regions defined by the US Department of Health and Human Services (“HHS regions”). We fit Poisson regression models to the weekly state-level counts of reported deaths due to P&I from January 5, 2015 to February 8, 2020 (see Supplement for details). The baseline was then projected forward until March 28, 2020. We adjusted for seasonality using harmonic variables and included year-to-year baseline variation. Influenza activity was controlled for by adjusting for the percent of tests that were positive for influenza in the previous week. The 1-week lag between the testing data and the mortality data accounts for the delay influenza testing and death. The number of all-cause deaths was used as a denominator. Poisson 95% prediction intervals were estimated by sampling from the uncertainty distributions for the estimated model parameters.^14^

To calculate the number of COVID-19 related excess deaths, we subtracted the expected number of deaths in each week from the observed number of deaths for the period February 9, 2020 to March 28, 2020. Because reporting of deaths for recent weeks is incomplete, NCHS calculates a ‘completeness’ score (between 0 and 1) based on the number of death reports that have been received from a state and the number expected from that state based on previous years. The excess deaths were divided by the completeness score for each week to get an estimate for excess cases, adjusted for reporting delays.

To evaluate the sensitivity of the estimates of excess mortality to the adjustment for influenza activity, we refit the regression excluding the influenza covariate. We also fit a model with the same structure to all-cause mortality (without a denominator). To get a measurement of outpatient visits related to COVID-19, the same model was fit to data on influenza-like illness.

### Code and data availability

The analyses were run using R v 3.6.1. All analysis scripts and archives of the data are available from https://github.com/weinbergerlab/excess_pi_covid

## RESULTS

Many states experienced a notable increase in the proportion of total deaths due to P&I starting in mid-March through March 28 compared to what would be expected based on the time of year and influenza activity (**Figure 1, Figure 2**). Expressed as the relative increase above the baseline, these increases were particularly notable in New Jersey, Washington, New York, Illinois, and Georgia. The increase in New York was largely driven by spikes in New York City (**Figure S1**).

**Figure 1.**
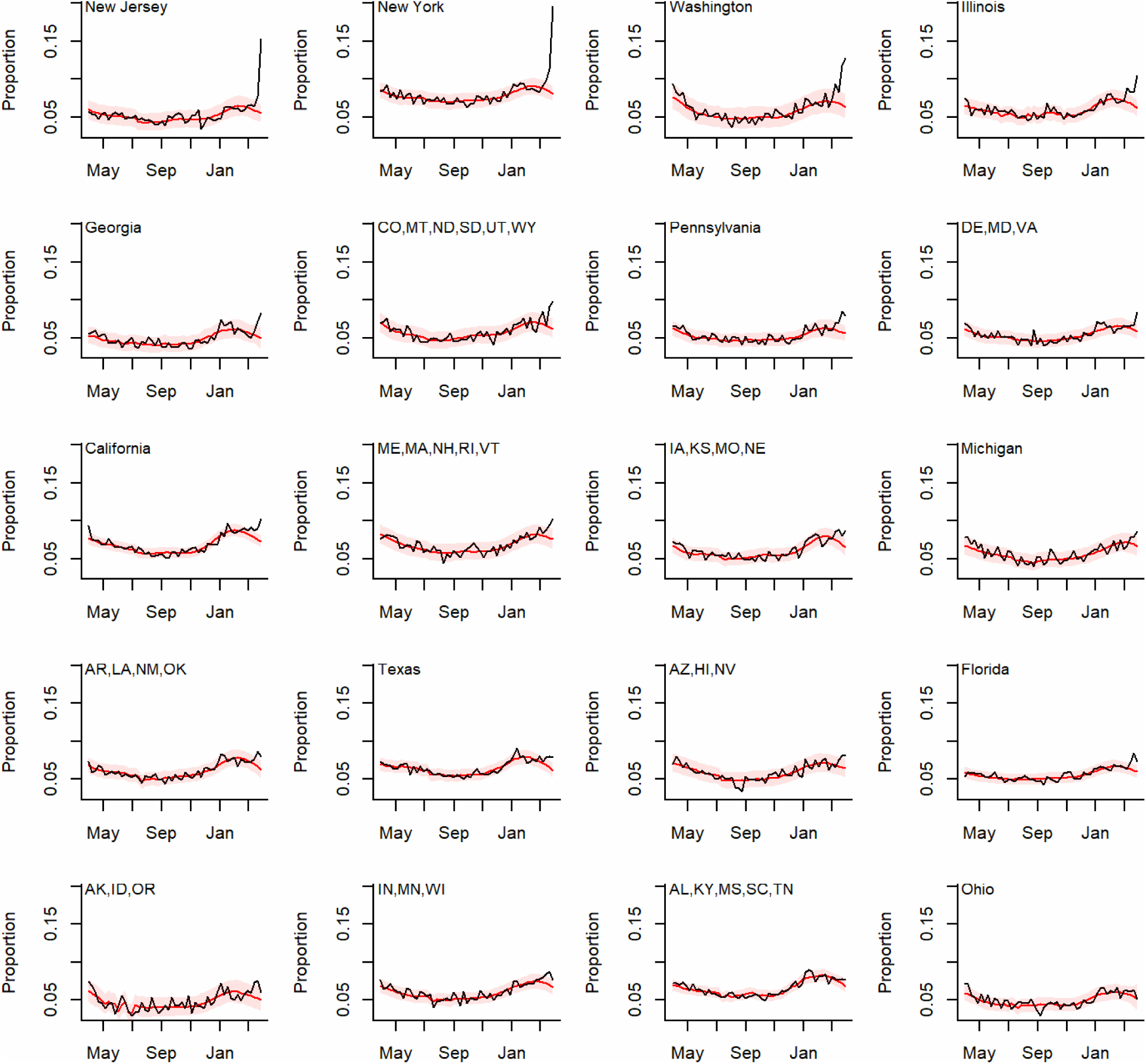
Proportion of deaths in each week that were due to pneumonia and influenza. The black line shows the observed proportion, and the red line and shaded region show the seasonal baseline (adjusted for influenza activity) +/- 95% prediction intervals. States are ordered based on the size of the relative increase over baseline at the last time point.

**Figure 2.**
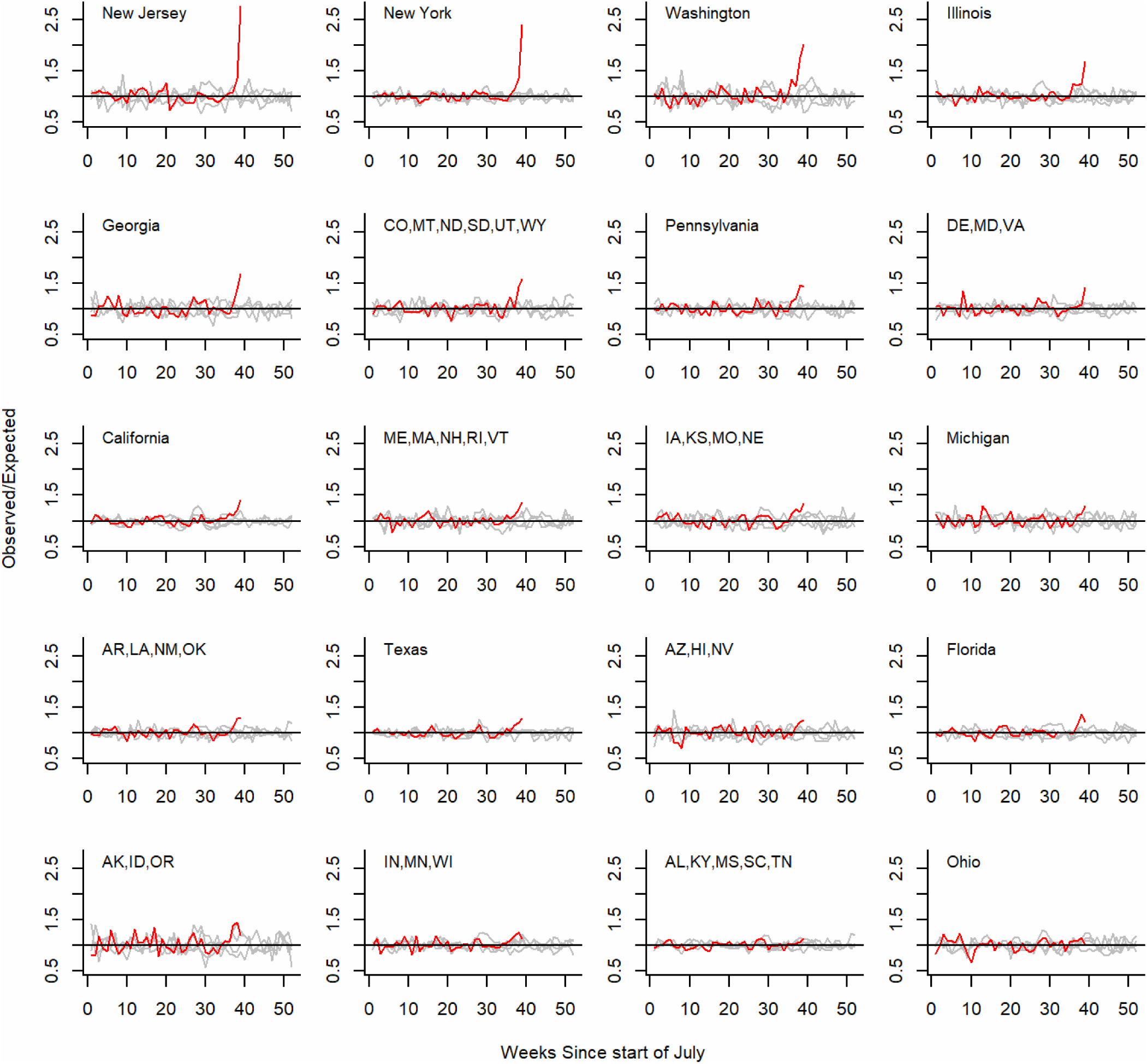
Observed/expected deaths due to pneumonia and influenza in each week for 2019-20 (red) compared with the same period of time in the past 4 years (grey). Values greater than 1 indicate that there were more deaths than expected due to pneumonia & influenza in that week.

In some states, such as Florida and Georgia, the increase in deaths due to P&I preceded the widespread adoption of testing for the novel coronavirus by several weeks (**Figure 3**). As a result, the increase in P&I preceded the first reported COVID-19 deaths, and the excess P&I was greater than the number of reported COVID-19 deaths each week. In contrast, in Washington state, there was concordance between reported COVID-19 deaths, excess P&I deaths, and the increase in testing.

**Figure 3.**
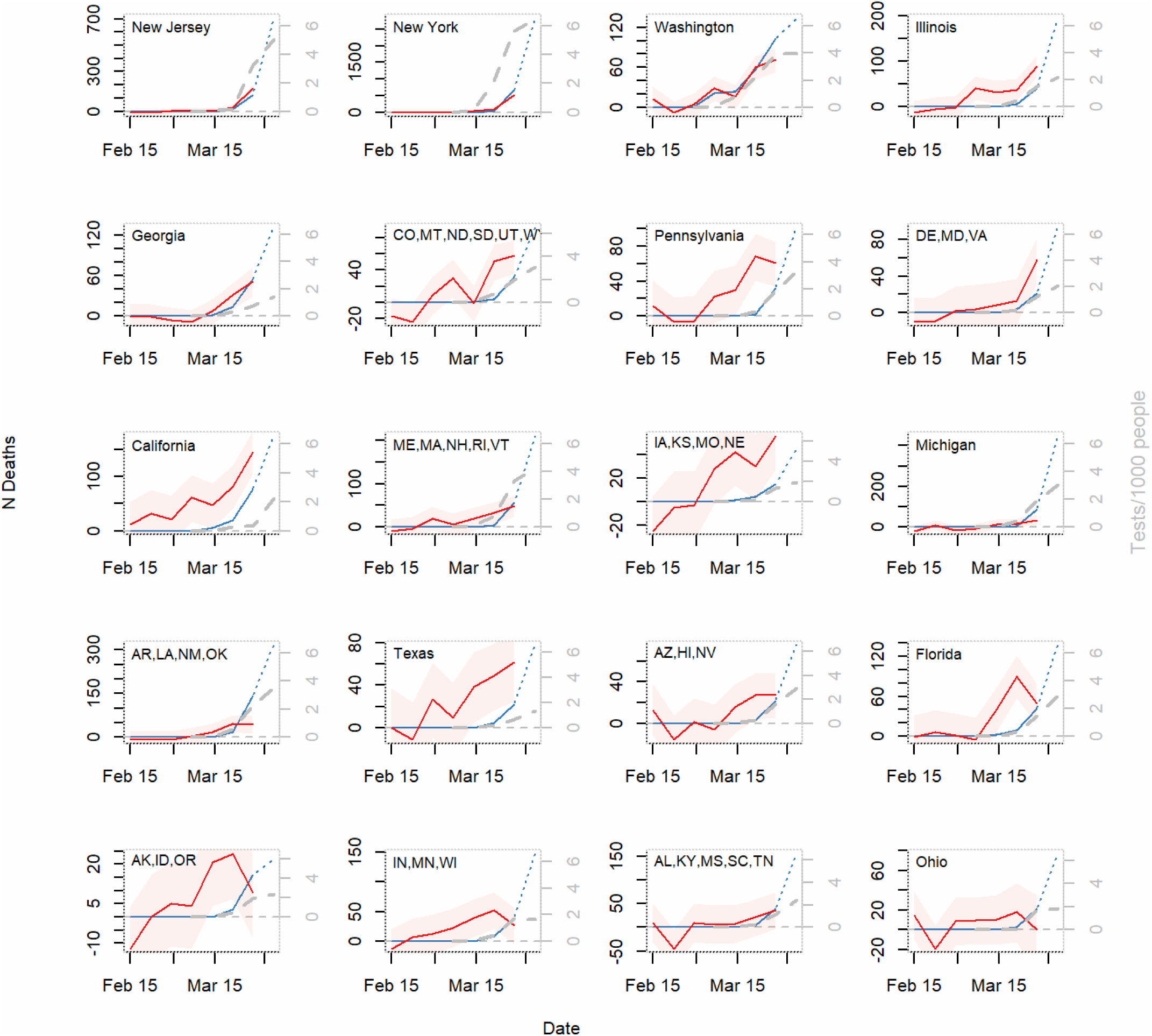
Trends in excess mortality due to pneumonia and influenza (red) compared with trends in reported deaths due to COVID-19 by week and state for February 9, 2020 to March 28, 2020. The dashed blue line represents reported COVID-19 deaths in weeks for which the pneumonia and influenza data were not yet available. The thick dashed gray line shows the volume of tests performed/1000 people in that week.

The timing of the epidemic varied across states. Some states and regions that have not yet seen large increases in deaths are earlier in the epidemic, though there is some indication of sharp increases in outpatient visits for ILI (eg, Ohio, Texas **Figure 4)**. The increase in P&I mortality typically lags behind the increase in ILI visits, so increases in excess P&I deaths might be expected in coming weeks.

**Figure 4.**
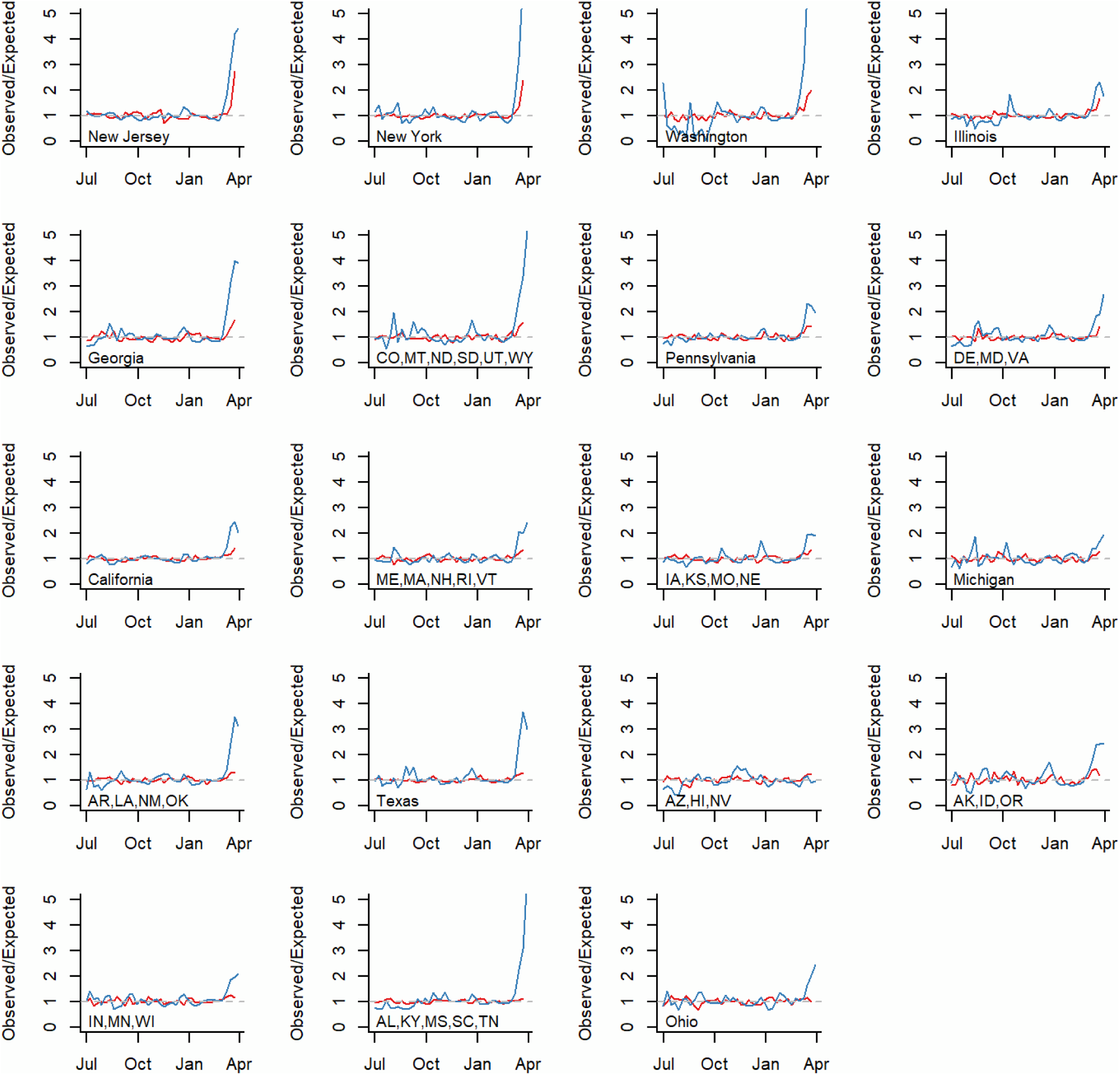
Unexplained increases in deaths due to pneumonia/influenza (red) compared with unexplained increases of cases of influenza-like illness at outpatient providers (blue) from July 2019-March 22, 2020.

The degree of under-reporting in some states can be seen when comparing the estimated excess deaths due to P&I with the number of reported COVID-19 cases through March 28 (**Table 1, Figure S4**). For instance, California had 101 reported deaths due to COVID-19 and 399 (286, 512) excess P&I deaths. Likewise, Texas had 176 (86, 266) excess P&I deaths and 27 reported deaths through March 28 (**Table 1)**. In contrast, for New York and Washington, there was agreement between the reported COVID-19 deaths and the excess P&I deaths (**Figure 3, Figure S1)**. In Michigan, the number of excess P&I deaths was notably lower than the number of reported COVID-19 deaths, potentially reflecting reporting delays in the P&I data. Overall, in the states we evaluated for the period from February 9 to March 28, 2020 there were 3101 (2769, 3433) excess P&I deaths, compared with 1958 reported deaths due to COVID-19 from the COVID Tracking Project. There were 2537 deaths due to COVID-19 provisionally reported by NCHS (for the entire U.S.) during this period.

**Table 1.** Excess deaths due to pneumonia and influenza and deaths due to COVID-19, as reported by the COVID Tracking Project from February 9, 2020 through Mar 28, 2020.

| State | Total P&I deaths | Excess P&I deaths | Reported COVID-19 deaths | Excess P&I deaths/1,000,000 people |
| --- | --- | --- | --- | --- |
| NY | 2654 | 620(526, 714) | 728 | 31.9 (27.1, 36.7) |
| WA | 736 | 187(139, 235) | 207 | 24.6 (18.2, 30.9) |
| NJ | 860 | 206(154, 258) | 140 | 23.2 (17.4, 29) |
| IL | 1266 | 185(117, 253) | 47 | 14.6 (9.2, 20) |
| PA | 1237 | 182(115, 249) | 34 | 14.2 (9, 19.4) |
| CA | 3428 | 399(286, 512) | 101 | 10.1 (7.2, 13) |
| ME,MA,NH,RI,VT | 1258 | 114(44, 183) | 59 | 10.1 (3.9, 16.3) |
| IA,KS,MO,NE | 1498 | 124(47, 200) | 20 | 8.7 (3.3, 14.2) |
| CO,MT,ND,SD,UT,WY | 953 | 105(45, 165) | 36 | 8.6 (3.7, 13.5) |
| FL | 2039 | 180(93, 267) | 54 | 8.4 (4.3, 12.4) |
| IN,MN,WI | 1795 | 150(65, 235) | 49 | 8.2 (3.6, 12.9) |
| AK,ID,OR | 466 | 51(10, 93) | 19 | 7.6 (1.4, 13.8) |
| AR,LA,NM,OK | 1402 | 95(21, 169) | 158 | 6.9 (1.5, 12.3) |
| GA | 677 | 68(18, 118) | 69 | 6.4 (1.7, 11.2) |
| TX | 2105 | 176(86, 266) | 27 | 6.1 (3, 9.2) |
| AZ,HI,NV | 994 | 62(0, 125) | 25 | 5.3 (0, 10.6) |
| DE,MD,VA | 1188 | 67(-3, 136) | 25 | 4.3 (-0.2, 8.7) |
| OH | 973 | 44(-19, 106) | 25 | 3.7 (-1.6, 9.1) |
| MI | 979 | 37(-27, 101) | 92 | 3.7 (-2.7, 10.1) |
| AL,KY,MS,SC,TN | 2661 | 47(-58, 152) | 43 | 1.9 (-2.4, 6.2) |

Deaths due to P&I represent just a fraction of all deaths caused by COVID-19. To highlight this, we compared increases in deaths due to any cause in New York and New Jersey with increases in deaths due to P&I and reported deaths due to COVID-19 (**Figure S2)**. There were 2-3 times as many excess all-cause deaths as reported COVID-19 deaths or excess P&I deaths. In New York City, this discrepancy was even more stark, with 3-4 times as many excess all-cause deaths as P&I deaths (**Figure S3)**. The observation that excess P&I deaths underestimate the burden of COVID-19 is born out in the NCHS data, where fewer than half of the deaths recorded as being due to COVID-19 had pneumonia listed as a cause of death.^13^ The proportion of COVID-19-coded deaths with a pneumonia diagnosis itself varies considerably by state (**Table S2**).

In sensitivity analyses, we refit the seasonal baseline but did not adjust for influenza activity (**Table S3**), and we calculated excess P&I mortality without using a correction factor for incomplete reporting (**Table S4)**. These changes influence the specific estimates for some of the states with smaller increases but do not affect the overall pattern.

Mortality data are released weekly, and regularly-updated analyses are available at https://weinbergerlab.github.io/excess_pi_covid/.

## DISCUSSION

Excess P&I mortality has been used as a method for tracking influenza mortality for more than a century. Here we used a similar strategy to capture COVID-19 deaths that had not been attributed specifically to the pandemic coronavirus. Monitoring trends in broad mortality outcomes, like P&I or all-causes, provides a window into the magnitude of the mortality burden missed in official tallies of COVID-19 deaths. Given the lack of adequate testing and geographical variability in testing intensity, this type of monitoring provides key information on the severity of the epidemic in different geographic regions. It also provides some indication of the degree to which viral testing is missing deaths associated with COVID-19 directly or indirectly. Our findings suggest that the degree to which deaths due to COVID-19 are being correctly attributed to SARS-CoV-2 varies by state. Some states, such as Florida and Pennsylvania, might have missed deaths early on and might be undercounting deaths by a substantial degree currently. Other states, like Washington, have an accurate estimate of the mortality burden of the pandemic virus due to intense testing. And in states that have been hit hard by the pandemic virus, such as New Jersey or New York, the total excess mortality burden is 2-3 times that ascribed to COVID-19 in official statistics. Together, these findings demonstrate that estimates of the death toll of COVID-19 based on excess P&I and all-cause mortality will be more reliable than those relying only on reported deaths, particularly in places that lack widespread testing.

Local epidemics of COVID-19 started at different times across the US. States with early epidemics, like New York and New Jersey, are now reporting large increases in both P&I and all-cause mortality. Other states that had later epidemic onsets are just now entering the period of rapid growth of deaths. Because some states instituted social distancing measures at an earlier phase of the epidemic, they might benefit more from reducing the intensity of the peak in deaths.

Syndromic endpoints, such as deaths due to P&I, outpatient visits for ILI, and emergency department visits for fever, can provide a crude but informative measure of the progression of the outbreak.^15^ These measures themselves can be biased by changes in health seeking behavior and how conditions are recorded. However, in the absence of widespread and systematic testing for COVID-19, they provide a useful measure of epidemic progression and the effect of interventions.

To get a complete picture of the burden of the burden of deaths due to COVID-19, it will be necessary to evaluate spikes in all-cause mortality, as we have done for New York and New Jersey here. However, it is difficult to do such analyses reliably in real time with provisional death statistics because the data are incomplete for recent weeks, and the delays in reporting can only be determined retrospectively. Our analyses here suggest that excess P&I deaths represent a fraction of all of the deaths related to COVID-19 (25-50% based on preliminary data), so the P&I excess mortality estimates we present here represent a lower bound of the burden.

There is often a lag in the reporting of death statistics. NCHS reports data from 2 weeks prior, but these provisional data are typically still incomplete. In our main analysis, we addressed this by adjusting deaths due to P&I with all-cause deaths, with the assumption that reporting delays for P&I are similar to reporting delays for other causes of death. When calculating excess deaths, we attempted to adjust for reporting delays by using the NCHS estimate of data completeness as a multiplier. However, this estimate of data completeness is itself based on numbers of deaths during previous years and might not be reliable during the pandemic period. If anything, we would expect that longer reporting lags would tend to underestimate the excess mortality burden of COVID-19 in real-time analyses.

It is estimated that the COVID-19 epidemic in Europe predates that in the US by a few weeks. Several European countries have experienced a high death toll, particularly Italy and Spain. Consolidated excess mortality estimates are not yet available in these countries, although real-time all-cause mortality data from the EuroMomo project (https://www.euromomo.eu/) offers a glimpse of the total burden of the epidemic. Many European countries have experienced sharp increases in all-cause deaths in recent weeks, primarily in individuals 65 yrs and over, and also in 15-64 yrs

In conclusion, monitoring syndromic causes of death can provide crucial additional information on the severity and progression of the COVID-19 pandemic. Estimates of excess deaths due to P&I and all-causes will be less biased by variations in viral testing, but reporting lags have to be properly accounted for. Together with information on official tallies of COVID-19 deaths, monitoring excess mortality provides a key tool in evaluating the effects of an ongoing pandemic.

## Data Availability

All analysis scripts and archives of the data are available from https://github.com/weinbergerlab/excess_pi_covid

## ACKNOWLEDGEMENTS

The authors thank Jenny Chen for assistance in preparing references for the manuscript.

## Supplement to

### SUPPLEMENTAL FIGURES

**Figure S1:**
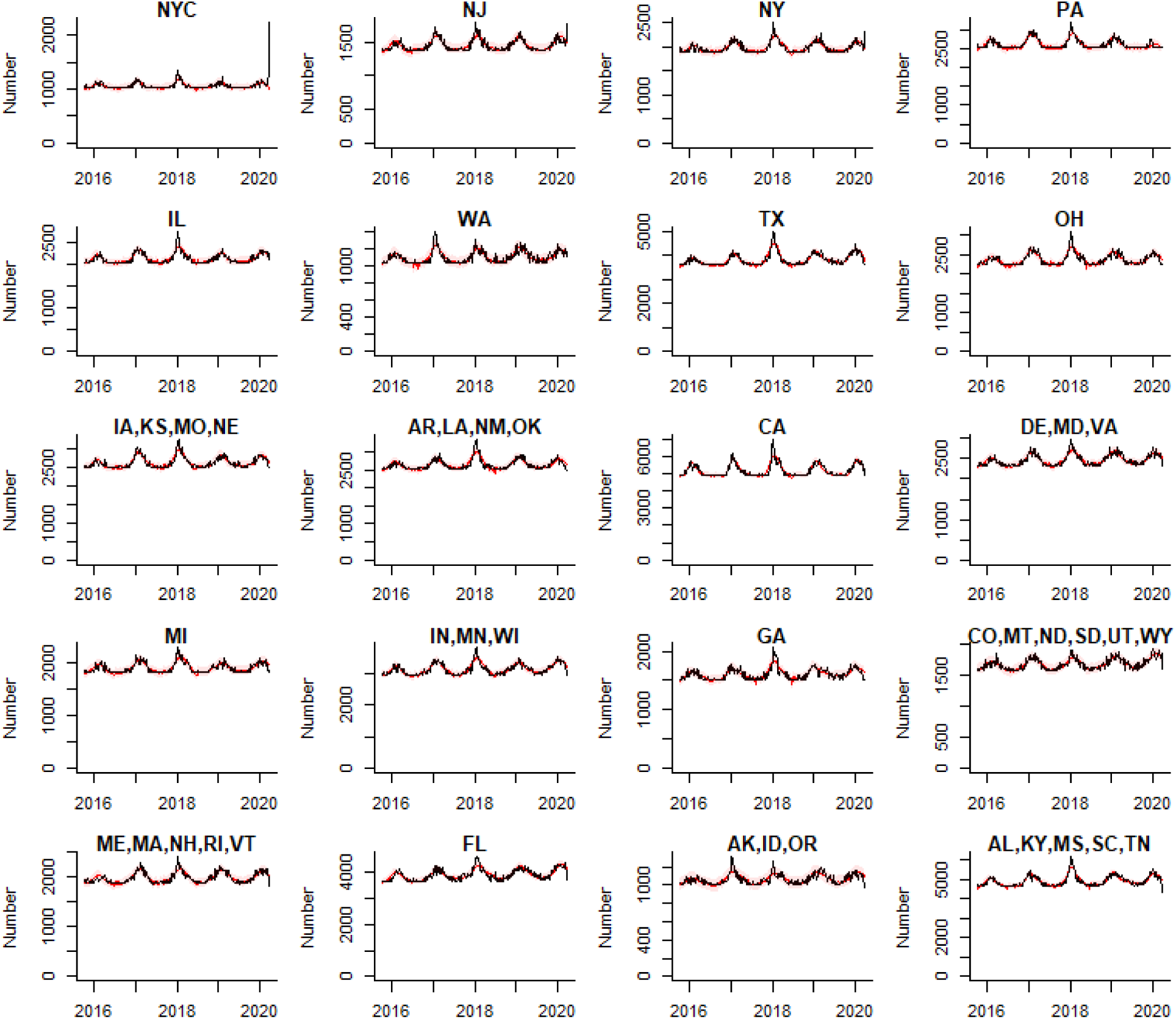
Excess all-cause deaths. The black line shows the observed number of *all* deaths per week, regardless of cause. The red line and shaded area represent the 95% Prediction Interval. The latest data is for the week ending 2020-03-28. Note that these are adjusted for percent completeness of the data using the NCHS’ estimate of data completeness. There are clear jumps in all-cause mortality in NY and NJ, other states are stable or decreasing, likely due to reporting delays.

**Figure S2:**
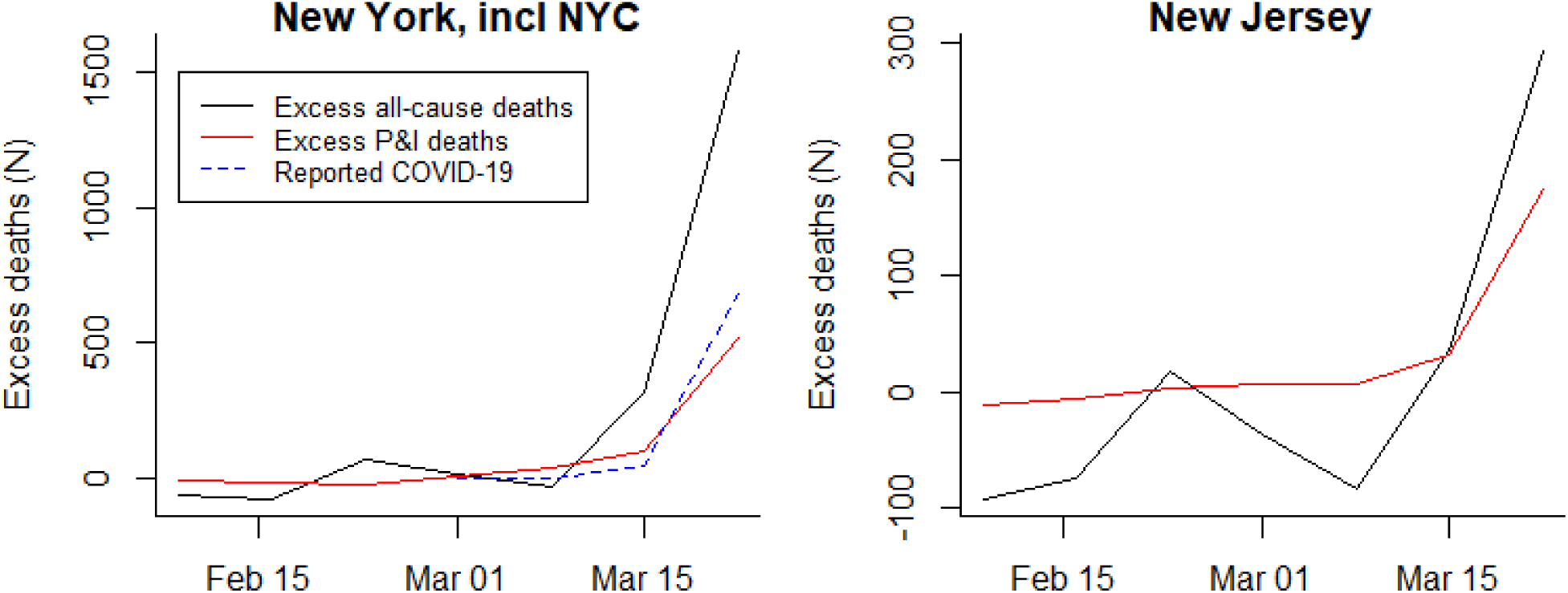
Excess all-cause deaths (black) vs Excess deaths due to pneumonia and influenza (red) and reported COVID-19 deaths from the COVID Tracking Project (blue dashed line) For New York (including New York City) and New Jersey.

**Figure S3.**
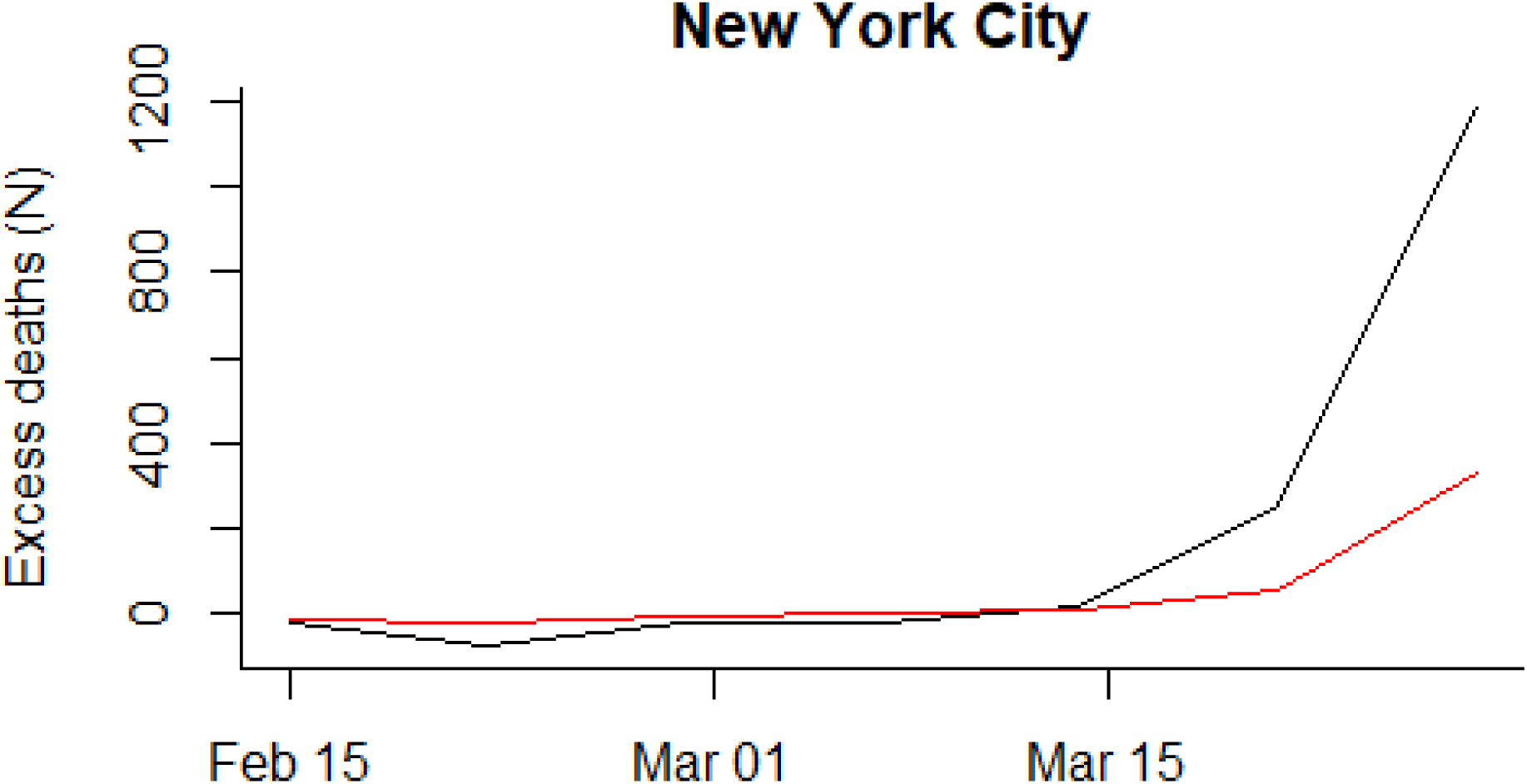
Excess P&I deaths per week (red) vs all-cause excess deaths (black) in New York City only.

**Figure S4.**
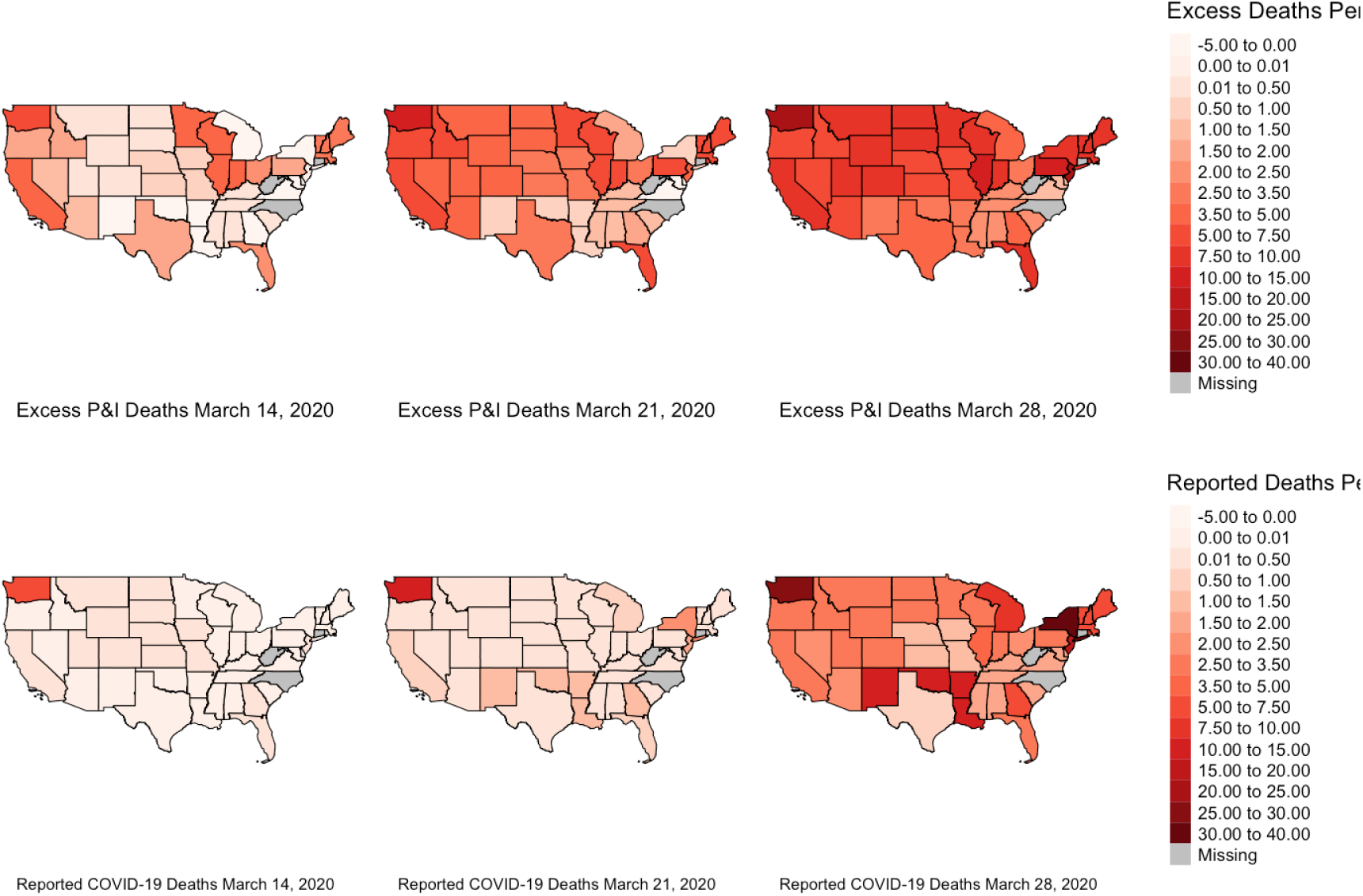
Map of Excess deaths by state and COVID-19 deaths reported by The COVID Tracking Project

### SUPPLEMENTAL TABLES

**Table S1:** Comparison of data sources: Observed and Excess deaths due to pneumonia & influenza, and COVID-19, from February 9, 2020 through Mar 28, 2020

| **Week ending** | **Total P&I deaths** | **Excess P&I deaths** | **NCHS Reported COVID-19 Deaths, with pneumonia code** | **NCHS Reported COVID-19 Deaths** | **All reported COVID-19 deaths (covidtracking.com)** |
| --- | --- | --- | --- | --- | --- |
| 2020-02-15 | 3971 | -71(-202, 60) | 0 | 0 | 0 |
| 2020-02-22 | 3867 | -132(-262, -2) | 0 | 0 | 0 |
| 2020-02-29 | 3998 | 67(-62, 196) | 3 | 5 | 1 |
| 2020-03-07 | 4092 | 270(143, 397) | 11 | 19 | 22 |
| 2020-03-14 | 4046 | 455(332, 578) | 22 | 44 | 36 |
| 2020-03-21 | 4317 | 887(767, 1007) | 200 | 435 | 229 |
| 2020-03-28 | 4877 | 1622(1505, 1739) | 928 | 2034 | 1670 |

**Table S2:** Proportion of COVID-19 deaths with a pneumonia code, by state, through most recent date. Note these values will be greater than those in other tables, which are 2 weeks behind

| State | NCHS Reported COVID-19 Deaths | NCHS Reported COVID-19 Deaths, with pneumonia code | Proportion with pneumonia |
| --- | --- | --- | --- |
| Total US | 6930 | 3161 | 0.46 |
| Alabama | 17 | 6 | 0.35 |
| Alaska | 1 | 1 | 1 |
| Arizona | 26 | 13 | 0.5 |
| Arkansas | 10 | 2 | 0.2 |
| California | 187 | 102 | 0.55 |
| Colorado | 107 | 60 | 0.56 |
| Delaware | 2 | 2 | 1 |
| District of Columbia | 5 | 5 | 1 |
| Florida | 157 | 93 | 0.59 |
| Georgia | 74 | 42 | 0.57 |
| Hawaii | 1 | 0 | 0 |
| Idaho | 13 | 4 | 0.31 |
| Illinois | 77 | 47 | 0.61 |
| Indiana | 43 | 19 | 0.44 |
| Iowa | 12 | 2 | 0.17 |
| Kansas | 9 | 3 | 0.33 |
| Kentucky | 6 | 3 | 0.5 |
| Louisiana | 154 | 64 | 0.42 |
| Maine | 10 | 5 | 0.5 |
| Maryland | 75 | 42 | 0.56 |
| Massachusetts | 172 | 85 | 0.49 |
| Michigan | 309 | 142 | 0.46 |
| Minnesota | 17 | 6 | 0.35 |
| Mississippi | 34 | 17 | 0.5 |
| Missouri | 22 | 9 | 0.41 |
| Montana | 3 | 1 | 0.33 |
| Nebraska | 3 | 2 | 0.67 |
| Nevada | 27 | 21 | 0.78 |
| New Hampshire | 15 | 8 | 0.53 |
| New Jersey | 593 | 303 | 0.51 |
| New Mexico | 1 | 0 | 0 |
| New York | 1130 | 650 | 0.58 |
| New York City | 2862 | 1065 | 0.37 |
| North Dakota | 1 | 0 | 0 |
| Ohio | 8 | 4 | 0.5 |
| Oklahoma | 17 | 5 | 0.29 |
| Oregon | 21 | 13 | 0.62 |
| Pennsylvania | 220 | 98 | 0.45 |
| Rhode Island | 1 | 1 | 1 |
| South Carolina | 36 | 13 | 0.36 |
| South Dakota | 4 | 1 | 0.25 |
| Tennessee | 25 | 11 | 0.44 |
| Texas | 47 | 18 | 0.38 |
| Utah | 8 | 3 | 0.38 |
| Vermont | 17 | 6 | 0.35 |
| Virginia | 39 | 15 | 0.38 |
| Washington | 278 | 142 | 0.51 |
| Wisconsin | 34 | 7 | 0.21 |
| Puerto Rico | 18 | 13 | 0.72 |

**Table S3.** Comparison of baselines that are or are not adjusted for influenza. Observed and Excess deaths due to pneumonia & influenza, and COVID-19, from February 9, 2020 through March 28, 2020.

| State | Excess P&I deaths (unadjusted) | Excess P&I deaths (adjusted) | Reported COVID-19 deaths |
| --- | --- | --- | --- |
| NY | 620(526, 714) | 620(526, 714) | 728 |
| CA | 393(280, 506) | 399(287, 512) | 101 |
| NJ | 206(154, 258) | 206(154, 258) | 140 |
| WA | 187(139, 235) | 187(139, 235) | 207 |
| IL | 185(117, 253) | 185(117, 253) | 47 |
| FL | 180(93, 267) | 180(93, 267) | 54 |
| TX | 159(71, 247) | 176(88, 264) | 27 |
| PA | 156(96, 216) | 182(122, 242) | 34 |
| IN,MN,WI | 146(62, 230) | 150(66, 234) | 49 |
| ME,MA,NH,RI,VT | 110(40, 180) | 114(44, 183) | 59 |
| IA,KS,MO,NE | 110(35, 185) | 124(48, 199) | 20 |
| CO,MT,ND,SD,UT,WY | 101(41, 161) | 105(46, 165) | 36 |
| AR,LA,NM,OK | 74(3, 145) | 95(24, 166) | 158 |
| DE,MD,VA | 65(-4, 134) | 67(-2, 136) | 25 |
| AZ,HI,NV | 61(-1, 123) | 62(0, 125) | 25 |
| GA | 47(-2, 96) | 68(20, 117) | 69 |
| AK,ID,OR | 45(5, 85) | 51(11, 92) | 19 |
| AL,KY,MS,SC,TN | 38(-65, 141) | 47(-57, 150) | 43 |
| MI | 37(-27, 101) | 37(-27, 101) | 92 |
| OH | 34(-23, 91) | 44(-14, 101) | 25 |

**Table S4.** Comparison of estimates when the estimates for excess cases are adjusted based on the estimated completeness of the database. Excess deaths due to pneumonia & influenza from February 9, 2020 through Mar 28, 2020 with or without adjustment for delayed reporting

| state | Excess P&I (unadjusted) | Excess P&I (adjusted for reporting delay) |
| --- | --- | --- |
| NY | 514(420, 608) | 620(526, 714) |
| CA | 370(257, 482) | 399(287, 512) |
| NJ | 206(154, 258) | 206(154, 258) |
| WA | 156(108, 204) | 187(139, 235) |
| IL | 165(97, 233) | 185(117, 253) |
| PA | 75(15, 135) | 182(122, 242) |
| FL | 179(92, 266) | 180(93, 267) |
| TX | 88(0, 177) | 176(88, 264) |
| IN,MN,WI | 101(17, 185) | 150(66, 234) |
| IA,KS,MO,NE | 62(-13, 137) | 124(48, 199) |
| ME,MA,NH,RI,VT | 61(-9, 130) | 114(44, 183) |
| CO,MT,ND,SD,UT,WY | 53(-7, 113) | 105(46, 165) |
| AR,LA,NM,OK | 67(-5, 138) | 95(24, 166) |
| GA | 40(-9, 88) | 68(20, 117) |
| DE,MD,VA | 39(-30, 108) | 67(-2, 136) |
| AZ,HI,NV | 46(-16, 109) | 62(0, 125) |
| AK,ID,OR | 17(-23, 58) | 51(11, 92) |
| AL,KY,MS,SC,TN | 42(-61, 145) | 47(-57, 150) |
| OH | 13(-44, 70) | 44(-14, 101) |
| MI | 46(-18, 110) | 37(-27, 101) |

### SUPPLEMENTAL METHODS

#### Statistical model

We developed a regression model for P&I deaths in epidemiological year *i* (July-June) and week *t* as a function of seasonal parameters and the percent positive influenza tests in the prior week. Models were fit separately for each location with data from January 5, 2015 to February 8, 2020; fitted values were projected for the period until March 28, 2020. Let PI_Deaths_i,t_ be the number of P&I deaths and let Flu_Pct_Pos_i,t-1_ be the percent positive influenza tests. We modeled

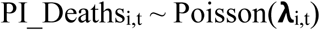

where

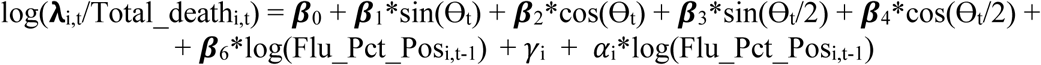

and

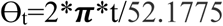

To compute prediction intervals, we used the following procedure. Once the regression coefficients were estimated, we extracted the estimated asymptotic covariance matrix for the parameters and constructed a multivariate normal distribution approximating the sampling distribution, centered at the estimated parameter values. We drew 100 samples from this parameter distribution, computed the resulting mean value **λ**_i,t_, and then drew 100 samples from the Poisson distribution with this mean. This resulted in 10,000 samples from an empirical predictive distribution of PI_Deaths_i,t_. Empirical 95% prediction intervals were computed by taking the 2.5th and 97.5th percentiles of this resulting distribution. In further sensitivity analyses, we evaluated a model in which Flu_Pct_Pos was excluded altogether.

## Notes

### Competing Interest Statement

DMW has received consulting fees from Pfizer, Merck, GSK, and Affinivax for topics unrelated to this manuscript and is Principal Investigator on a research grant from Pfizer on an unrelated topic. VEP has received reimbursement from Merck and Pfizer for travel expenses to Scientific Input Engagements unrelated to the topic of this manuscript. All other authors report no relevant conflicts.

### Funding Statement

DMW acknowledges support from grants R01AI123208 and R01AI137093 from the National Institute of Allergy and Infectious Diseases/National Institutes of Health. VEP acknowledges support from grants R01AI112970 and R01AI137093 from the National Institute of Allergy and Infectious Diseases/National Institutes of Health. NGR acknowledges support from grant R35GM119582 from the National Institute of General Medical Sciences/National Institutes of Health and 1U01IP001122 from the Centers for Disease Control and Prevention (CDC). TC acknowledges support from R01AI146555 from the National Institute of Allergy and Infectious Diseases/National Institutes of Health. The content is solely the responsibility of the authors and does not necessarily represent the official views of the National Institutes of Health, New York City Department of Health and Mental Hygiene or CDC. LS acknowledges support from the Carlsberg Foundation grant # CF20-0046. FWC acknowledges support from NICHD grant 1DP2HD091799-01.

